# Hemodialysis Patients Show a Highly Diminished Antibody Response after COVID-19 mRNA Vaccination Compared to Healthy Controls

**DOI:** 10.1101/2021.03.26.21254259

**Authors:** Benedikt Simon, Harald Rubey, Andreas Treipl, Martin Gromann, Boris Hemedi, Sonja Zehetmayer, Bernhard Kirsch

## Abstract

1.

**Background and Objectives:** Hemodialysis patients are prone to infection with SARS-COV2 and show a high probability of a severe course of disease and high mortality when infected. In many countries hemodialysis patients are prioritised in vaccination programs to protect this vulnerable community. However, no hemodialysis patients were included in efficacy trials of SARS CoV-2 vaccines and therefore efficacy and safety data for this patient group are lacking. These data would be critical, since hemodialysis patients showed decreased responses against various other vaccines and this could mean decreased response to SARS CoV-2 vaccines.

**Design, setting, participants, and measurements:** We conducted a prospective cohort study consisting of a group of 81 hemodialysis patients and 80 healthy controls who were vaccinated with mRNA vaccine BNT162b2 (BionTech/Pfizer, 2 doses with an interval of 21 days). Anti-SARS-COV-2 S antibody response in all participants was measured 21 days after the second dose. The groups were compared with univariate quantile regressions and a multiple analysis. Adverse events (AEs) of the vaccination were assessed with a standardized questionnaire. We also performed a correlation of HBs-Antibody response with the SARS-COV-2 antibody response in the hemodialysis patients.

**Results:** Dialysis patients had significantly lower Anti-SARS-COV-2 S antibody titres than healthy control patients 21 days after vaccination with BNT162b2 (median dialysis Patients 171 U/ml versus median controls 2500 U/ml). Age also had a significant but less pronounced influence on antibody titres. Dialysis patients showed less AEs than the control group. No significant correlation was found for Hepatitis B vaccine antibody response and SARS CoV-2 vaccine antibody response.

**Conclusions:** Hemodialysis patients exhibit highly diminished SARS-COV-2 S antibody titres compared to a cohort of controls. Therefore these patients could be much less protected by SARS CoV-2 mRNA vaccination than expected. Alternative vaccination schemes must be considered and preventive measures must be maintained after vaccination.

## 1. Introduction

The SARS COV-2 pandemic has had and continues to have a profound impact on daily lives and all aspects of medicine[1–3]. Patients undergoing dialysis on a regular basis are especially prone to infection with the virus due to unavoidable exposure (frequent transports to and from dialysis centres and procedures there[4,5]) and severe course of disease with a 28-day-mortality of up to 16-35%[5–7]. Consequentially, in most countries dialysis patients are prioritized to receive vaccines against COVID-19.

One of the first vaccines approved worldwide, BNT162b2 (Comirnaty™, Pfizer/Biontech), was shown to elicit in study participants a measurable antibody response which correlated with protection from severe disease[8]. A two dose vaccination regimen reduced severe disease by over 90%[8]. However, no end stage renal disease patients were included in this study and data of efficacy and safety of this patient group is therefore lacking. These data would be critical since dialysis patients showed decreased response against various other vaccines (e.g., Hepatitis B[9], Pneumococcus[10] or Influenza vaccines[11]). Decisions like vaccine schedule and dosing depend on such data. Therefore, the aim of this study is to measure antibody response in dialysis patients and compare them to the antibody response of a control group of healthy volunteers (healthcare workers). In addition, we will compare the adverse events in both cohorts and analyse whether a correlation between humoral response to Hepatitis B vaccine and response to BNT62b2 exists.

## 2. Material and Methods

We conducted a prospective cohort study design to elucidate the antibody response of vaccination with BNT162b2 in dialysis patients vs. healthy controls.

### 2.1 Study Population

Haemodialysis Patients were considered eligible if they were on dialysis for at least 3 months and had received vaccination with mRNA vaccine BNT162b2 (BionTech/Pfizer Comirnaty, 2 doses administered with an interval of 21 days according to national vaccination schedule). The participants of the healthy control group consisted of volunteer health care workers who had been vaccinated using the same regimen. Participants in both groups needed to be 18-99 years old. Pregnant women and individuals with known COVID19 infection in the past (diagnosed via patient history and serological test for nucleocapsid (N) antibody, see 2.3 Serological assessment) were excluded from the study. The study protocol was approved by the local Ethics committee and participants were enrolled after written informed consent was obtained. 92 dialysis patients were initially recruited for the study; 3 patients were excluded after testing positive for antibodies against SARS CoV-2 N protein. 8 patients were excluded (5 because conditions did not allow for vaccination within 21 days, 2 deaths, 1 transplantation). Finally, 81 dialysis patients were included in the study, 23 women and 58 men. Mean age was 67 years (median 70 years, age range 34 to 86 years).

81 volunteer healthcare workers were initially recruited for the study; one person was excluded due to a positive N antibody test. Finally, 80 healthy controls were included in the study, 50 women and 30 men. Mean age was 49 years (median 52 years, age range 29 to 65 years). All demographic data are summarized in Table 1.

**Table 1.** Demographics of the study population. COPD, Chronic obstructive pulmonary disease. RAAS, Renin-Angiotensin-Aldosteron system. CNI, Calcineurin inhibitors. MMF, Mycophenolat-Mofetil. EPO, Erythropoetin.

|  | Dialysis patients<br>(n=81) | Control<br>group (n=80) |
| --- | --- | --- |
| **Age (y, mean, range)** | 67 (34-86) | 49 (29-65) |
| **Men** | 71 (55%) | 30 (24%) |
| **Risk factors** |  |  |
| Diabetes | 31 | 2 |
| COPD | 23 | 0 |
| Hypertension | 68 | 21 |
| **Primary Kidney Disease** |  |  |
| Diabetes | 25 | n.a. |
| Vascular disease | 27 | n.a. |
| Glomerulonephritis | 10 | n.a. |
| unknown | 1 | n.a. |
| other | 18 | n.a. |
| **Medication** |  |  |
| RAAS-Inhibitors usage | 35 | 0 |
| Immunosuppressants usage (Steroids, CNI, MMF) | 9 | 2 |
| Vitamin D supplements usage | 61 | 0 |
| EPO usage | 74 | 0 |

Medical histories of dialysis patients were extracted from medical records. Medical histories of the control group were assessed by a standardized questionnaire.

### 2.2 Processing of blood samples

Blood draws were performed at 3 weeks after the second administration of BNT62b2. Samples were centrifuged on a Hettich Rotanta 460r centrifuge at 3000rpm for 10 minutes, aliquoted and anonymized. They were then stored at -70° C and thawed prior to testing.

### 2.3 Serological assessment

All samples were analysed with an Elecsys®Anti-SARS-CoV-2 test, measuring the nucleocapsid (N) antibodies. A positive result in this test lead to an exclusion from the study due to a high probability of a clinical or subclinical COVID19 infection in the past[12].

The antibody response elicited by vaccination with BNT62b2 was measured using an Elecsys ® Anti-SARS-CoV-2 S on a Cobas e 801 platform according to specifications[13]. Results were recorded as ranging from 0 (<=0.40 U/ml, lower limit of detection (LOD)) to 2500 (>=2500 U/ml, upper LOD) and assigned to anonymized patient data.

For the correlation of Hepatitis B vaccination responders with the Sars-COV2 vaccination responders we defined the following cut-offs: Hepatitis B vaccine responders were defined as hepatitis B antibody Titer > 20 I.U./ml after at least one completed Hepatits-B vaccination cycle (3 doses with Engerix B 40µg).

### 2.4 Adverse events

Adverse events (AE) of the vaccination were assessed separately for both vaccine doses and both groups with a standardized questionnaire.

AEs were divided into two categories: local AEs (pain at injection site, redness and/or swelling at injection site and induration at injection site) and systemic AEs (fatigue, headache, muscle and/or joint pain, fever, gastrointestinal symptoms [diarrhoea, nausea, vomiting] or other AEs).

Patients were asked to grade their AE after both vaccine doses in subjective severity. Grading was performed on a scale from 1 to 4. Grade 1 AE signified mild (does not interfere with activity); Grade 2 moderate, (interferes with activity); Grade 3 severe, (prevents daily activity); and Grade 4 (emergency department visit or hospitalization), in analogy to the FDA toxicity grading scale[14].

### 2.5 Statistics

To analyse the influence of group (dialysis patients versus controls), sex, and age on titre, univariate quantile (median) regressions were performed. Because of imbalances in sex and age between the two groups, a multiple analysis was computed for all variables which were significant in the univariate analysis. The quantile regression was chosen due to the skewed distribution of titre (many patients with maximum titre observation of 2500). Bootstrap was applied to construct standard errors and perform statistical tests for each independent factor (5000 replications), median and interquartile range (IQR) were given to compare the groups descriptively.

A further quantile regression was performed for the variable hepatitis, which is only available for the dialysis group. In case of a significant result, additionally age and sex were considered.

The significance level was set to 0.05. The analyses were performed with R 4.0.2 and the quantreg package.[15]

Boxplots were generated using Prism (GraphPad, 2021 version; Prism -GraphPad). AE bar graphs were created in Excel 2019 (Microsoft, included in Office 365; Microsoft Excel, Tabellenkalkulationssoftware | Microsoft 365).

## 3. Results

81 dialysis patients and 80 control group patients were tested for their antibody response after receiving two doses of BNT162b2 (BionTech/Pfizer, “Comirnaty™”). Their characteristics, risk factors and other data are shown in Table 1.

Anti-SARS-CoV2 antibody titres were measured 3 weeks after the second vaccine dose was administered in both groups. The univariate analysis shows that dialysis patients have a highly significant lower titre than the control group (median dialysis Patients 171 U/ml, IQR: 477.7, versus median controls 2500 U/ml, IQR 943.5). Gender and age also have a significant influence on titre (see table 2). Note that the variables group and gender are confounded: in the control group 62.5% are women, whereas in the dialysis group only 28.4% are female. In our study cohort men have a lower median titre (median for men of 367.5 U/ml, IQR=1650, versus 1542 U/ml for women, IQR: 1790) than women. Age has a negative influence on titre; with increasing age, the titre decreases on average. The Spearman correlation coefficient for the two variables is -0.62.

**Table 2.**
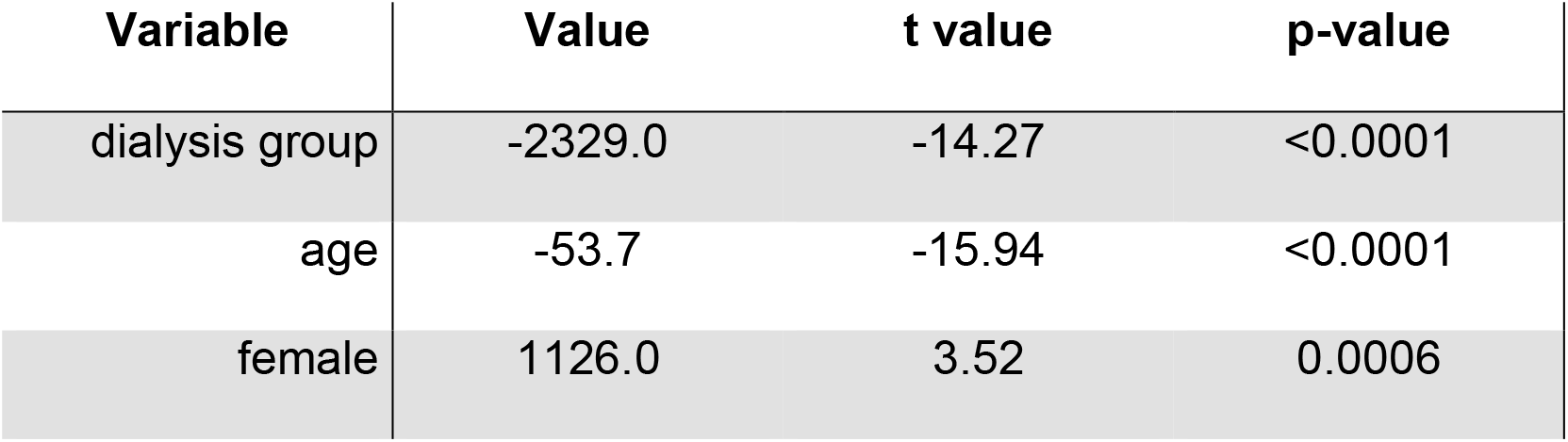
Univariate analysis of variables’ influence on antibody titre calculated using univariate quantile regressions. P values <0,05 were considered significant.

**Table 3.** Multiple analysis of variables’ influence on antibody titre calculated using univariate quantile regressions. P values <0,05 were considered significant.

| Variable | Value | t value | p-value |
| --- | --- | --- | --- |
| dialysis group | -1998.1 | -10.27 | <0.0001 |
| age | -9.4 | -2.57 | 0.011 |
| female | 17.4 | 0.17 | 0.86 |

The multiple analysis results in a highly significant influence of group – dialysis patients have significantly lower titres than controls -and a smaller influence of age on titre (Table 2). Sex has no significant influence anymore in the multiple analysis.

Adverse event (AE) reports were analysed and compared descriptively between the two groups. No Grade 4 (emergency department visit or hospitalization) AEs were reported in either group. The control group reported more local AEs after both vaccine doses (first dose (56 vs. 33 AEs), second dose (66 vs 17 AEs)) and also more systemic AEs after both vaccine doses (first dose (30 vs 7 AEs) and second dose (51 vs 7 AEs) compared to the dialysis group. See **Fehler! Verweisquelle konnte nicht gefunden werden**. Figure 2 and Figure 3 for a graphical representation.

**Figure 1.**
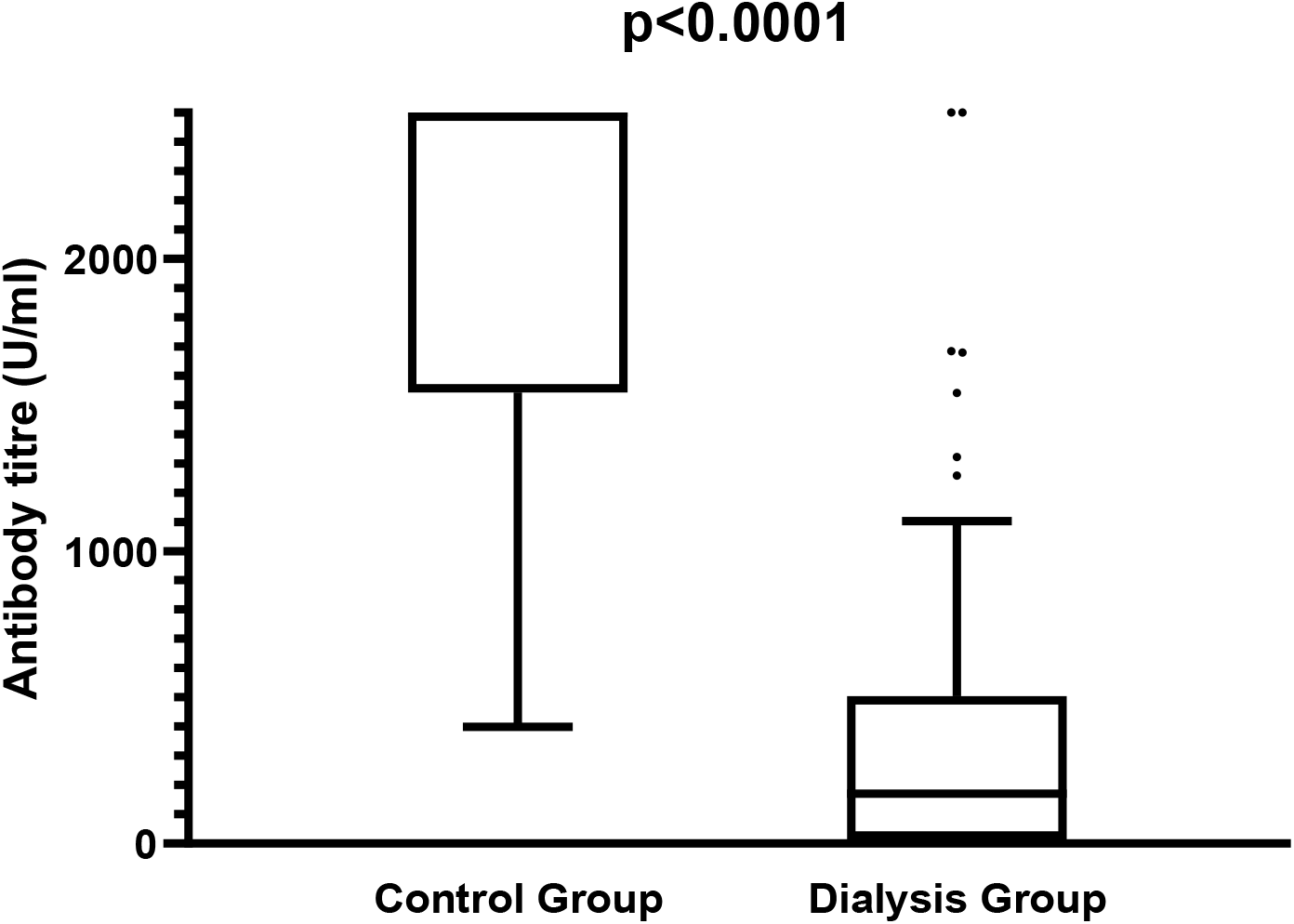
Boxplot of SARS CoV-2 specific antibody titres (controls vs dialysis patients) 21 days after the 2nd vaccine dose. Note that the maximum titre in the test system used is 2500 U/ml (cut-off). Control group titres are significantly higher than dialysis group titres (p<0.0001)

**Figure 2.**
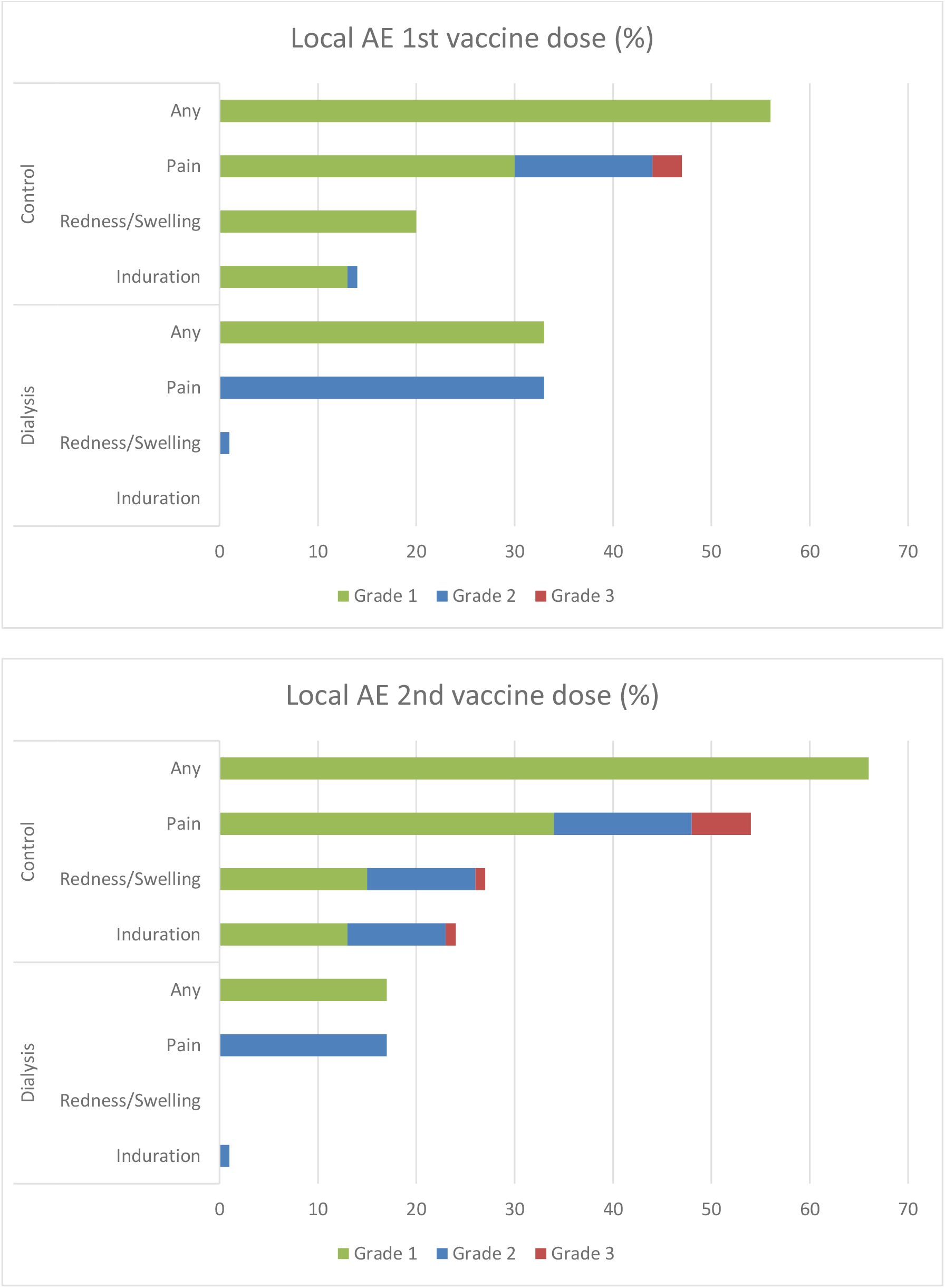
local adverse events after vaccination with BNT162b2. All numbers are percentages of dialysis (n=81) and control (n=80) patients. AEs were recorded via a standardized questionnaire and graded by patients (Grade 1 mild, does not interfere with activity; Grade 2 moderate, interferes with activity; Grade 3 severe, prevents daily activity. No Grade 4 events (emergency visit or hospitalisation) were reported).

**Figure 3.**
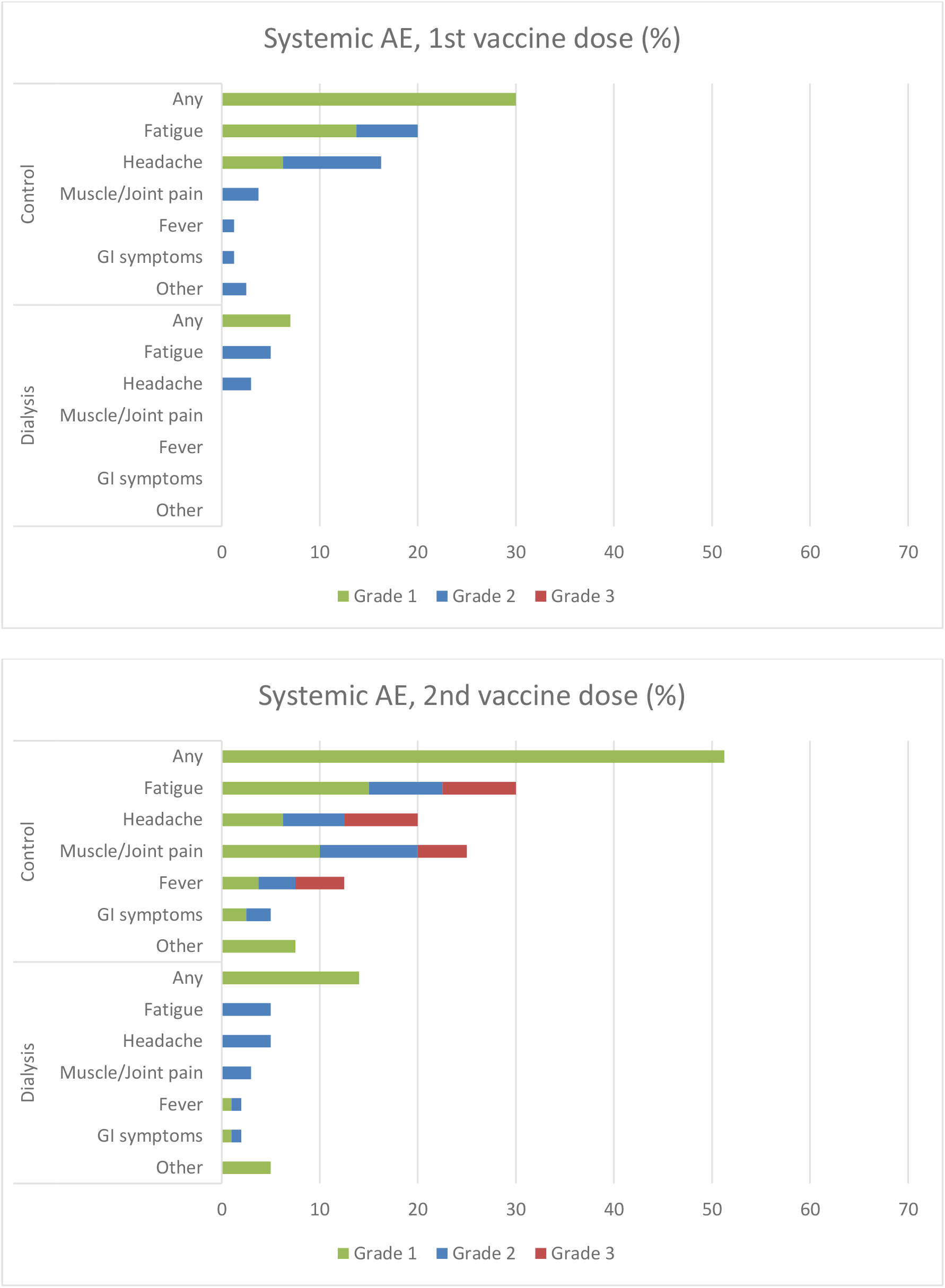
systemic adverse events after vaccination with BNT162b2. All numbers are percentages of dialysis (n=81) and control (n=80) patients. AEs were recorded via a standardized questionnaire and graded by patients (Grade 1 mild, does not interfere with activity; Grade 2 moderate, interferes with activity; Grade 3 severe, prevents daily activity. No Grade 4 events (emergency visit or hospitalisation) were reported). GI, gastrointestinal AEs (diarrhoea, nausea, vomiting).

In the dialysis group, patients with an antibody response >20 I.U./ml to hepatitis B vaccine (“responders”) have a higher SARS CoV-2 antibody titre (responders: median=223.5, IQR=587; non-responders: median=159, IQR=450). However, this difference is not significant in the quantile regression (value: -50, t-value: -0.37, p=0.71). Note that the sample size for this analysis is only 81 patients.

## 4. Discussion

Antibody titres after diverse vaccinations tend to be considerably lower in dialysis patients, with a greater percentage of said patients lacking measurable titres using a conventional vaccination scheme compared to healthy patients, e.g. Hepatitis B vaccine[16],[9], Pneumococcus[10] or Influenza vaccines[11]. Therefore, it is uncertain whether vaccinating against SARS COV-2 in this patient collective will result in sufficient immune response and, by consequence, protection against infection. To our knowledge, this is the first study describing anti-S antibody titres in dialysis patients vs. healthy controls.

In the control group, antibody titres 3 weeks after two doses of BNT162b2 were significantly higher than in the dialysis group (p<0.0001). All patients in the control group had a titre greater than 200U/ml, signifying a robust antibody response. On the other hand, in the dialysis group 43 patients (53%) had an antibody titre lower than 200 U/ml (signifying neutralization below maximum), 22 patients (27%) had a titre lower than 29 U/ml (likely no neutralization) and seven patients (9%) had no detectable antibodies at all. This signifies a weaker antibody response in dialysis patients overall, making them less likely to be able to neutralize SARS CoV-2 virus even after two doses of vaccine. Since dialysis patients are more exposed to infection[4,5] and prone to a severe course of disease this could pose a grievous problem in this vulnerable community.

The possibility of a delayed response is remote, but it exists, so longitudinal follow-up studies are warranted, especially since it was shown that antibody levels after natural COVID-19 infection seem to decline in hemodialysis patients over time[17] and the same could happen after vaccination.

The gold standard to measure neutralizing capacity of patient serum antibodies is a plaque reduction neutralization test[18], where cells are incubated with virus and the dilution in which virus growth is inhibited is measured. A plaque assay was also used in efficacy studies for BNT162b2 (Pfizer/BionTech) COVID-19 vaccine as a surrogate marker for protection from severe disease[8].

The test used in our study, the Elecsys ® Anti-SARS-CoV-2 S, is by nature not a neutralisation assay; it uses a recombinant protein representing the receptor binding domain (RBD) of the SARS COV-2 S (spike) protein as an antigen and quantitatively measures antibodies directed against this protein. Still, recent data indicate a good correlation with a direct virus neutralization test and a surrogate neutralization assay (GenScript® cPass™ SARS-CoV-2 Neutralization Antibody Detection Kit)[19]: Briefly, the serum of all patients who achieved an antibody titre of 29 U/ml or above in the Elecsys test system correlate with a 1:5 titre in neutralization assays and therefore possess some measure of neutralizing capacity. Sera with an Elecsys antibody result of equal to or greater than 200 U/ml correlate with maximal neutralizing capacity in the neutralization assays.[20]

The Elecsys test was also used to quantify samples from the WHO International Standard and Reference Panel for anti-SARS-CoV-2 antibody[21] and showed good correlation.[20]

Therefore, we feel comfortable using this test as a robust and accepted surrogate marker for an immune response achieved by vaccination.

The time point of testing three weeks after the second dose was chosen because the system used in this study measures IgM as well as IgG. In analogy to natural infection, after three weeks the initial IgM boost response should have subsided in most patients, while the IgG response is at its peak[22]. Thus, in theory this is the optimal time point to measure protective, durable IgG responses.

Adverse events (AE) were reported significantly more frequent and with higher grading by the control group than the dialysis group. This could mean a more noticeable immune reaction in control group patients. Due to the relatively small sample-size, our study does not aim to detect statistical differences of adverse events between groups. Therefore, further studies are needed to uncover potential causal relationship between occurrence of AE and immune response in dialysis patients.

No statistically significant correlation was found in the dialysis patients between responders to Hepatitis B vaccination (defined as HBs-Antibody Titer >20 IU/ml after at least one completed vaccination cycle) and response to BNT162b2. This could reflect different immune mechanisms and levels of reactogenicity in response to the two vaccines. The basis for Hepatitis B vaccine non-response has not yet been elucidated; probably multifactorial in origin, at least some part can safely be attributed to immune suppression of dialysis-patients (low antibody response seems to correlate with the degree of renal failure[23]). Possible explanations include immune cell disturbances[24–26]. Also gene variations in vaccine response genes (e.g., Interferon-λ4 polymorphisms) were shown to influence Hepatitis B vaccine response in dialysis patients[27].

Age and gender were distributed unequally between the control and dialysis group: While in the control group, the majority of patients were female, in the dialysis group males predominated. Patients of the control group were on average younger than patients in the dialysis group. To account for these confounding factors, we performed a multiple analysis. We found that dialysis vs. control group showed the highest impact on the antibody-titre, while age influenced the antibody titres to a much lesser extent. Gender was not a significant factor in the multiple analysis, showing that its significance in the univariant analysis was a consequence of group composition.

One limitation of our study is that the clinical significance of even a plaque-based neutralization assay has not been widely tested; it is probable, but not yet proven, that high antibody titres in our test system and, by correlation, in neutralization assays protect patients from severe infection courses. Further studies are needed to validate the impact of protective antibody titres in clinical settings.

Another limitation in this regard is that our test system only tested humoral (antibody), but no cellular (T-cell) immune response. Since the correlates of protection in SARS CoV-2 infection remain unknown as of this date, the cellular part of the adaptive immune system probably plays a role in protection from COVID-19[28] which is not reflected in our investigation.

In summary, our data show that the hemodialysis patients in our study, who are at a very high risk for infection with COVID-19 and severe course of disease and mortality[5–7,29], developed an antibody response which is significantly lower than the antibody response in our control group. This finding has implications for preventative measures beyond vaccination (masks, social distancing and hand hygiene, testing strategies, patient isolation etc.) which need to be maintained for protection. Further studies for alternative vaccination strategies (dosing, schedule) are urgently needed.

## Data Availability

All data included in manuscript

## References

1. Dubey S, Biswas P, Ghosh R et al. Psychosocial impact of COVID -19. Diabetes Metab Syndr 2020; 14: 779–788

2. Nicola M, Alsafi Z, Sohrabi C et al. The socio-economic implications of the coronavirus pandemic (COVID-19): A review. Int J Surg 2020; 78: 185 –193

3. Wastnedge EAN, Reynolds RM, van Boeckel SR et al. Pregnancy and COVID-19. Physiol Rev 2021; 101: 303–318

4. ÖGN. SARS-CoV-2 Infektionen im ÖDTR, Stand 12.02.2021. Österreichische Gesellschaft für Nephrologie. 2020 Dec 12.

5. Francis A, Baigent C, Ikizler TA, Cockwell P, Jha V. The urgent need to vaccinate dialysis patients against severe acute respiratory syndrome coronavirus 2: a call to action. Kidney Int 2021; 99: 791 –793

6. Hilbrands LB, Duivenvoorden R, Vart P et al. COVID-19-related mortality in kidney transplant and dialysis patients: results of the ERACODA collaboration. Nephrol Dial Transplant 2020; 35: 1973–1983

7. Jager KJ, Kramer A, Chesnaye NC et al. Results from the ERA-EDTA Registry indicate a high mortality due to COVID -19 in dialysis patients and kidney transplant recipients across Europe. Kidney Int 2020; 98: 1540 –1548

8. Sahin U, Muik A, Derhovanessian E et al. COVID-19 vaccine BNT162b1 elicits human antibody and T(H)1 T cell responses. Nature 2020; 586: 594 –599

9. Stevens CE, Alter HJ, Taylor PE, Zang EA, Harley EJ, Szmuness W. Hepatitis B vaccine in patients receiving hemodialysis. Immunogenicity and efficacy. N Engl J Med 1984; 311: 496–501

10. Fuchshuber A, Kühnemund O, Keuth B, Lütticken R, Michalk D, Querfeld U. Pneumococcal vaccine in children and young adults with chronic renal disease. Nephrol Dial Transplant 1996; 11: 468–473

11. Scharpé J, Peetermans WE, Vanwalleghem J et al. Immunogenicity of a standard trivalent influenza vaccine in patients on long -term hemodialysis: an open-label trial. Am J Kidney Dis 2009; 54: 77 –85

12. Cdc. Interim Guidelines for COVID -19 Antibody Testing [Internet]. 2020 [updated 2021 Mar 24; cited 2021 Mar 24]. Available from: https://www.cdc.gov/coronavirus/2019-ncov/lab/resources/antibody-tests-guidelines.html#anchor_1616005971325

13. Roche Diagnostics International Ltd. Elecsys Anti -SARS CoV2 -S factsheet [Internet]. 2020. Available from: https://diagnostics.roche.com/content/dam/diagnostics/Blueprint/en/pdf/cps/Elecsys-Anti-SARS-CoV-2-S-factsheet-SEPT-2020-2.pdf

14. Toxicity Grading Scale for Healthy Adult and Adolescent Volunteers Enrolled in Preventive Vaccine Clinical Trials. FDA. <time datetime=“2019-05-17T14:30:29Z”>Fri, 2019 May 17 - 14:30</time>.

15. R Core Team. R: A language and environment for statistical computing. [Internet]. R Foundation for Statistical Computing, Vienna, Austria. 2020

16. Edey M, Barraclough K, Johnson DW. Review article: Hepatitis B and dialysis. Nephrology (Carlton) 2010; 15: 137–145

17. Labriola L, Scohy A, Seghers F et al. A Longitudinal, 3 -Month Serologic Assessment of SARS-CoV-2 Infections in a Belgian Hemodialysis Facility. Clin J Am Soc Nephrol 2020

18. Muruato AE, Fontes-Garfias CR, Ren P et al. A high-throughput neutralizing antibody assay for COVID-19 diagnosis and vaccine evaluation. Nature Communications 2020; 11: 4059

19. Tan CW, Chia WN, Qin X et al. A SARS-CoV-2 surrogate virus neutralization test based on antibody-mediated blockage of ACE2 –spike protein–protein interaction. Nature Biotechnology 2020; 38: 1073–1078

20. Rubio-Acero R, Castelletti N, Fingerle V et al. In search for the SARS-CoV-2 protection correlate: A head-to-head comparison of two quantitative S1 assays in a group of pre-characterized oligo-/asymptomatic patients; 2021

21. WHO. WHO/BS.2020.2403 Establishment of the WHO International Standard and Reference Panel for anti -SARS-CoV-2 antibody [Internet]. 2020 [updated 2020 Nov 18]

22. Sun B, Feng Y, Mo X et al. Kinetics of SARS -CoV-2 specific IgM and IgG responses in COVID-19 patients. Emerg Microbes Infect 2020; 9: 940 –948

23. Kausz A, Pahari D. The value of vaccination in chronic kidney disease. Semin D ial 2004; 17: 9–11

24. Eleftheriadis T, Antoniadi G, Liakopoulos V, Kartsios C, Stefanidis I. Disturbances of acquired immunity in hemodialysis patients. Semin Dial 2007; 20: 440–451

25. Litjens NHR, Huisman M, van den Dorpel M, Betjes MGH. Impaired immune responses and antigen-specific memory CD4+ T cells in hemodialysis patients. J Am Soc Nephrol 2008; 19: 1483–1490

26. Agrawal S, Gollapudi P, Elahimehr R, Pahl MV, Vaziri ND. Effects of end -stage renal disease and haemodialysis on dendritic cell subsets a nd basal and LPS-stimulated cytokine production. Nephrol Dial Transplant 2010; 25: 737 –746

27. Grzegorzewska AE, Świderska MK, Marcinkowski W, Mostowska A, Jagodziński PP. Polymorphism rs368234815 of interferon -λ4 gene and generation of antibodies to hepatitis B virus surface antigen in extracorporeal dialysis patients. Expert Rev Vaccines 2020; 19: 293–303

28. Le Bert N, Tan AT, Kunasegaran K et al. SARS-CoV-2-specific T cell immunity in cases of COVID-19 and SARS, and uninfected controls. Nature 2020; 584: 457 –462

29. Windpessl M, Bruchfeld A, Anders H -J et al. COVID-19 vaccines and kidney disease. Nat Rev Nephrol 2021; 1–3

